# Long-term clinical, virological and immunological consequences of mpox virus infection or modified vaccinia virus Ankara vaccination: a 24-month prospective cohort study

**DOI:** 10.1101/2025.05.21.25327977

**Authors:** Christophe Van Dijck, Nicole Berens-Riha, Luca M. Zaeck, Stefanie Bracke, Jacob Verschueren, Jasmine Coppens, Fien Vanroye, Elisabeth Willems, Evi Bosman, Natalie De Cock, Bart Smekens, Leen Vandenhove, Odin Goovaerts, Anke Van Hul, Janne Wouters, Cécile Kremer, Bart Jacobs, Matilde Hens, Isabel Brosius, Elise De Vos, Eugene Bangwen, Sarah Houben, Pedro Henrique Lopes Ferreira Dantas, Patrick Soentjens, Emmanuel Bottieau, Chris Kenyon, Johan van Griensven, Thijs Reyniers, Niels Horst, Kevin K. Ariën, Marjan Van Esbroeck, Andrea Torneri, Koen Vercauteren, Wim Adriaensen, Rory D. de Vries, Joachim Mariën, Laurens Liesenborghs

**Author notes:** Corresponding author, Nationalestraat 155, 2000 Antwerp, Belgium. These authors contributed equally (Christophe Van Dijck, Nicole Berens-Riha, Luca M. Zaeck). These authors jointly supervised the work (Joachim Mariën, Laurens Liesenborghs).

## Abstract

**Background:** As mpox virus (MPXV) continues to circulate, a better understanding of the long-term clinical, virological, and immunological consequences of infection and MVA-BN vaccination is needed.

**Methods:** In thiscohort study, we prospectively followed MPXV-infected individuals and individuals vaccinated with MVA-BN up to 24 months. Mpox patients were assessed for physical and mental well-being. Saliva and anorectal swabs were tested by MPXV-PCR. Vaccinia virus (VACV) lysate and MPXV-E8 binding and MPXV-neutralising antibodies were analysed over time, taking into account the impact of childhood smallpox vaccination.

**Results:** We included 237 MPXV-infected individuals and 210 vaccinees; 93.9% (369/393) were gay or bisexual men who have sex with men, one-third (143/428, 33.4%) lived with HIV, and 31.8% (142/447) were born before or in 1976, (end of smallpox vaccination in Belgium). Scarring occurred in 31.7% (20/63) up to two years post-infection. MPXV was not detected ≥8 months post-infection. Without childhood smallpox vaccination, MVA-BN vaccinees had significantly lower binding antibody levels than MPXV-infected individuals (0.39-fold, 95%CI 0.27-0.55, *p*<.001 for VACV-lysate antibodies, and 0.60-fold, 95%CI 0.46-0.79, *p*<.001 for MPXV-E8 antibodies). MPXV-E8 antibody levels decreased most rapidly over time in smallpox-naïve MVA-BN vaccinees (−8%/month, 95%CI −9.09 to −7.42). MPXV-neutralising antibodies were rarely detected in smallpox-naïve MVA-BN vaccinees (estimated seroprevalence 3%, 95%CI 1-11%, *i.e.* 0.04-fold, 95%CI 0.01-0.12, *p*<.001 lower compared to smallpox-naïve MPXV-infected individuals).

**Discussion:** Lower antibody levels among MVA-BN vaccinees compared to MPXV-infected individuals suggest a less robust immunological priming induced by vaccination. Studies should assess if booster doses enhance the durability of the immunological memory.

**Funding:** Research Foundation–Flanders; Department of Economy, Science and Innovation Flanders; Netherlands Organization for Health Research and Development (ZonMw)

## Background

Mpox virus (MPXV), the causative agent of mpox, remains a pressing concern on the global health agenda as it continues to cause large international outbreaks.^1^ MPXV causes fever, lymphadenopathies, pustulating skin lesions, and complications such as proctitis, secondary bacterial infections, and scarring. In 2022, clade IIb MPXV spread globally, infecting over 85,000 individuals and disproportionately affecting gay, bisexual, and other men who have sex with men (GBMSM).^2^ By the end of 2022, the epidemic had subsided in many regions, including Europe, likely due to a combination of increased awareness, behavioural changes, and immunity conferred by infection and targeted vaccination campaigns.^3,4^ Nevertheless, sporadic clusters of clade IIb cases have continued to be reported and appear to have increased in frequency since mid-2023.^5^ Meanwhile, in 2023, a new lineage of MPXV, designated as subclade Ib, emerged in the Democratic Republic of the Congo, spreading internationally and resulting in over 20,000 confirmed cases of mpox.^6^ This sustained circulation of multiple MPXV clades highlights the urgent need to better understand the long-term health consequences of mpox and the durability of vaccine-induced immunity.

Studies performed within the first year post-vaccination with the modified vaccinia virus-Ankara Bavarian Nordic (MVA-BN) vaccine have demonstrated a rapid decline in antibody levels.^7–13^ Two-year follow-up data further indicate that antibody levels return to baseline over time.^14,15^ In contrast, antibody levels after MPXV-infection reach high levels in the first months post-infection,^7,16,17^ and orthopoxvirus-specific IgG and neutralising antibodies are maintained for at least one year in most patients.^18^ However, studies directly comparing large cohorts of vaccinated and infected individuals remain limited,^7^ and longitudinal data beyond 12 months post-infection are lacking.

Additionally, several case series and cohort studies have reported considerable morbidity associated with mpox-related scarring in the first six to 15 months post-infection.^19–23^ Yet, to our knowledge, only one study described scarring up to two years post-infection, and no studies have investigated longer-term viral persistence and psychological sequelae.

Here, we present data from mpox patients and MVA-BN vaccinees, up to 24 months post-infection or vaccination. We describe the long-term physical and mental health consequences of mpox, assess the presence of MPXV at various body sites after infection, and compare antibody dynamics between MPXV-infected and MVA-BN-vaccinated individuals.

## Methods

### Study Participants and Setting

This prospective cohort study (ClinicalTrials.gov: NCT05879965) took place at the Institute of Tropical Medicine (ITM) Antwerp, Belgium, a travel and sexual health clinic, where, since 2022, almost one-third of mpox patients in Belgium were diagnosed.^3,24^ In Belgium, MVA-BN was offered as post-exposure preventive vaccination from June 2022 onward, and as primary preventive vaccination to people at risk of infection from August until December, 2022 Vaccines were administered subcutaneously (SC, 0.5 mL per dose), except between September and November, 2022, when all vaccine doses were administered off-label intradermally (ID, 0.1 mL per dose) due to vaccine shortages. Smallpox vaccination with a live replicating vaccinia virus (VACV) based vaccine was routinely offered in Belgium up to 1976, so individuals born until that year likely received smallpox vaccine during childhood. Individuals without proof of smallpox vaccination history were offered two doses with a minimum interval of 28 days. Otherwise, only one dose of MVA-BN was offered.

### Study Design

Since May 2022, individuals diagnosed with acute MPXV infection have been included in a prospective clinical registry. In September 2022, the study protocol was amended to a prospective long-term cohort study, with follow-up visits at the clinic at approximately 1, 8, 16, and 24 months after diagnosis. In addition, the protocol was expanded to include individuals vaccinated with MVA-BN with study visits at 8, 16, and 24 months after their last vaccine dose. Besides this prospective collection of data and samples, the protocol allowed for retrospective inclusion of clinical record data and biobanked samples from individuals who had not been approached by the study team at the time of diagnosis. Interim results of this study have been reported previously.^19,24^ Mpox patients who received MVA-BN vaccination were followed up in the MPXV-infected cohort, not in the MVA-BN cohort.

### Data & sample collection

Among MPXV-infected individuals, biological samples (serum, anorectal swabs, saliva) were collected at each visit. Semen sampling (only at month 8) was optional. Data were collected on demographics, sexual behaviour, comorbidities, scarring, pain, fatigue and mental health. A standardised questionnaire was used to assess stigma related to scarring (Dermatology Life Quality Index, DLQI) at months 16 and 24. A 21-point mpox severity score was calculated based on the number of lesions (1-4 points), number of body areas involved (0-5 points), proctitis (3 points), urethritis (2 points), fever (2 points) and throat ache, lymphadenopathy, arthralgia or myalgia, headache, and fatigue (1 point, each; **Supplement p4**).^16^ Among MVA-BN vaccinees, serum samples were collected at each visit, along with data on demographics, sexual behaviour, and comorbidities.

### Laboratory analyses

Saliva, anorectal swabs and semen samples were analysed by real-time MPXV-PCR (**Supplement p2**). To assess the dynamics of the antibody response over time, we used three complementary serological assays, each targeting distinct aspects of the humoral immune response. First, an ELISA based on whole-cell VACV-lysate provided a broad measure of orthopoxvirus-reactive IgG responses.^25^ Second, a bead-based Luminex assay targeting MPXV surface protein E8 measured MPXV-specific IgG (**Supplement p2**). Third, an MPXV plaque reduction neutralisation test (PRNT) was used to evaluate the functional capacity of the antibody response in a random subset of samples.^25^ Neutralising antibodies were considered present if titres exceeded the 95^th^ percentile of titres observed in baseline (pre-mpox and pre-MVA-BN) serum samples from individuals born after the cessation of smallpox vaccination campaigns in 1976.

### Statistical analysis

Continuous variables were summarised as medians with interquartile range, categorical variables as counts with percentages. Logistic regression was used to examine associations between mpox-related scarring and key predictors: birth cohort (before or in 1976 versus after 1976), HIV infection, and mpox severity.

To evaluate antibody persistence from childhood smallpox vaccination, we compared baseline (pre-mpox and pre-MVA-BN) samples from participants born before or in 1976 versus after 1976. Binding antibody levels were log-transformed and analysed using linear regression. Estimates were back-transformed to reflect fold-differences in geometric means. MPXV-neutralising antibody prevalence was compared between birth cohorts using generalised linear models with a log-binomial distribution (GLM-lb) treating antibody positivity (titre above cutoff) as a binary outcome.

In an exploratory, nested case-control analysis among participants born before or in 1976, we investigated whether higher binding antibody levels and MPXV-neutralising antibody positivity in baseline samples – presumed to result from smallpox vaccination – were associated with protection against MPXV infection. We assumed that all participants were potentially exposed to MPXV during 2022, with cases defined as those who developed infection (MPXV-infected cohort), and controls as those who remained uninfected until at least August 2022 (MVA-BN vaccinees). Logistic regression was used to evaluate these associations.

To model long-term binding antibody dynamics from month 8 onward, we performed linear mixed-effects regression (LMER) with a random intercept for participants. To correct for the effect of childhood smallpox vaccination on antibody levels, participants were categorised into four sub-cohorts based on birth year (before or in 1976 vs after 1976) and exposure group (MPXV-infected vs MVA-BN vaccinated). Fixed effects included time (as a continuous variable), sub-cohort, and their interaction. Persistence of MPXV-neutralising antibodies was assessed using GLM-lb, restricted to a single post-month-8 observation per participant due to limited repeated measurements and low within-person variability.

Finally, associations between HIV-infection, mpox severity (continuous variable), or vaccination route (ID, SC, ID-ID, SC-SC, or mixed: ID-SC or SC-ID) and antibody levels and prevalence were individually evaluated using LMER or GLM-lb models. In case of non-convergence of the GLM-lb, logistic regression was used. These analyses were stratified by sub-cohort to address interaction effects and enhance interpretability.

Data were analysed with R (v4.4.2).

### Ethics

All participants provided written consent to the use of their data and samples. The study was approved by ITM’s Institutional Review Board (references 1628/22 and 1641/22) and the Ethics Committee of the University Hospital of Antwerp (UZA; references 3797 and 4981). The funders of the study had no role in study design, data collection, data analysis, data interpretation, or writing of the report.

## Results

### Participants

By January, 2025, 237 MPXV-infected individuals and 210 MVA-BN vaccinees were included (**Table 1**, **Figure 1**). The median age of the participants was 40 (IQR 33 to 48) years. The majority identified as men (425/443, 95.9%) and as GBMSM (369/393, 93.9%). One-third of participants (143/428, 33.4%) were living with HIV, of whom 16.7% (22/132) had a CD4 cell count <500 cells/mm³. Approximately one-third (142/447, 31.8%) of participants were born before or in 1976, and were presumed vaccinated against smallpox during childhood. About 40% of the participants (177/447, 39.6%) attended all three long-term follow-up visits.

**Figure 1:**
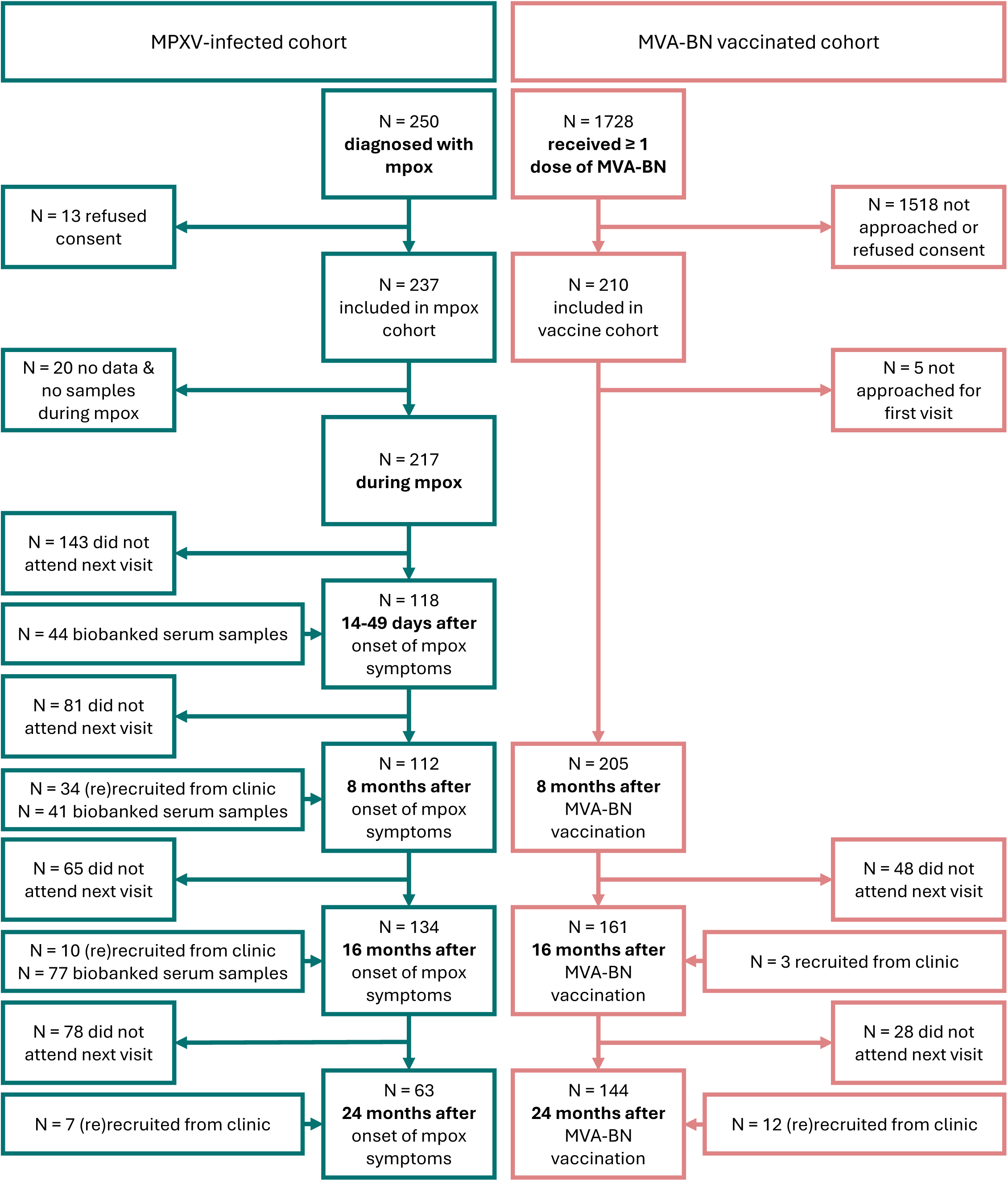
Participant flowchart. MPXV = mpox virus; MVA-BN = modified vaccinia virus Ankara – Bavarian Nordic

**Table 1:** Participant characteristics.

|  | MPXV-infected<br>(n=237) | MVA-BN vaccinated<br>(n=210) | Total (n=447) |
| --- | --- | --- | --- |
| **Age, median (IQR)** | 39 (33, 46) | 42 (34, 53) | 40 (33, 48) |
| Born ≤ 1976, n/N (%) | 62/237 (26.2) | 80/210 (38.1) | 142/447 (31.8) |
| Born > 1976, n/N (%) | 158/237 (73.8) | 130/210 (61.9) | 305/447 (68.2) |
| **Gender, n/N (%)** |  |  |  |
| Cis-man | 231/233 (99.1) | 194/210 (92.4) | 425/443 (95.9) |
| Trans-woman | 2/233 (0.85) | 4/210 (1.9) | 6/443 (1.4) |
| Cis-woman | 0/233 (0) | 5/210 (2.4) | 5/443 (1.1) |
| Non-binary/gender-fluid/gender-queer/other | 0/233 (0) | 7/210 (3.3) | 7/443 (1.6) |
| **Sexual orientation, n/N (%)** |  |  |  |
| Same sex | 171/184 (92.9) | 179/209 (85.6) | 350/393 (89.1) |
| Same & opposite sex | 4/184 (2.2) | 15/209 (7.2) | 19/393 (4.8) |
| Opposite sex | 9/184 (4.9) | 10/209 (4.8) | 19/393 (4.8) |
| Other | 0/184 (0) | 5/209 (2.4) | 5/393 (1.3) |
| **HIV, n/N (%)** |  |  |  |
| Living without HIV | 129/220 (58.6) | 155/208 (74.5) | 284/428 (66.4) |
| Using HIV-PrEP | 87/116 (75.0) | NA | - |
| Living with HIV | 91/220 (41.4) | 53/208 (25.5) | 143/428 (33.4) |
| CD4 < 500 | 13/80 (16.3) | 9/53 (17.0) | 22/132 (16.7) |
| **Immune suppression, n/N (%)** | 5/221 (2.3) * | 2/210 (0.95) ** | 7/431 (1.6) |
| **Splenectomy, n/N (%)** | NA | 3/202 (1.5) | NA |
| **Number of mpox lesions, n/N (%) <sup>£</sup>** |  | NA | NA |
| 0-4 | 88/214 (41.1) |  |  |
| 5-24 | 105/214 (49.1) |  |  |
| 25-100 | 19/214 (8.9) |  |  |
| >100 | 2/214 (0.93) |  |  |
| **Number of body areas<sup>§</sup> affected by mpox lesions, n/N (%)** |  | NA | NA |
| 0-1 | 84/195 (43.1) |  |  |
| 2-3 | 65/195 (33.3) |  |  |
| 4-5 | 28/195 (14.4) |  |  |
| >5 | 18/195 (9.2) |  |  |
| **Proctitis, n/N (%)** | 68/201 (33.8) | NA | NA |
| **Tonsillitis or throat ache, n/N (%)** | 24/186 (12.9) | NA | NA |
| **Urethritis, n/N (%)** | 14/201 (7.0) | NA | NA |
| **Lymphadenopathy, n/N (%)** |  | NA | NA |
| Regional, n/N (%) | 81/186 (43.5) |  |  |
| Generalised, n/N (%) | 9/186 (4.8) |  |  |
| **Fever or chills, n/N (%)** | 109/199 (54.8) | NA | NA |
| **Mpox severity score, median (IQR)<sup>#</sup>** | 8 (5.8, 10) | NA | NA |
| **Received MVA-BN vaccine, n/N (%)** | 28/237 (11.8) | 210/210 (100.0) | 238/447 (53.2) |
| Indication: Post-exposure preventive vaccination | 2/17 (11.8) | 10/204 (4.9) | 12/221 (5.4) |
| Indication: Primary preventive vaccination | 15/17 (88.2) | 194/204 (95.1) | 209/221 (94.6) |
| Received 1 dose | 9/28 (32.1) | 14/210 (6.7) | 23/238 (9.7) |
| Intradermally | 3/8 (37.5) | 4/14 (28.6) | 7/22 (3.2) |
| Subcutaneously | 5/8 (62.5) | 10/14 (71.4) | 15/22 (68.2) |
| Received 2 doses | 19/28 (67.9) | 196/210 (93.3) | 215/238 (90.3) |
| Intradermally – intradermally | 6/16 (37.5) | 117/196 (59.7) | 123/212 (58.0) |
| Intradermally – subcutaneously | 4/16 (25.0) | 39/196 (19.9) | 43/212 (20.3) |
| Subcutaneously – intradermally | 5/16 (31.3) | 8/196 (4.1) | 13/212 (6.1) |
| Subcutaneously – subcutaneously | 1/16 (6.3) | 32/196 (15.8) | 33/212 (15.6) |
| Vaccination interval among those who received 2 doses, median days (IQR) | 30 (28, 35) | 30 (28, 35) | 30 (28, 35) |

\*Three used immunosuppressive medication, one had diabetes mellitus type 1, one had HIV with a CD4-cell count of 250/mm<sup>3</sup>;
\*\*One had chronic renal insufficiency, one unknown.
<sup>£</sup> for 182/214 (85.0%) patients, this was the number of lesions present during consultation, estimated by the physician at diagnosis, for 32/214 (15.0%) patients, this was the maximum number of lesions during illness, reported by the participant at month 1
<sup>§</sup> evaluated body areas were head and neck; chest; abdomen; back; arms; hands; legs; feet; genital; anal
<sup>#</sup> calculation: see Supplementary Table 1
NA = data not available or not applicable

Among the mpox patients, 193/214 (90.2%) had fewer than 25 skin lesions, and in 149/195 (76.4%) of them, fewer than four body areas were affected (**Table 1**, **Figure 1**). About half of the patients reported fever (109/199, 54.8%), or lymphadenopathy (90/186, 48.4%), and 68/201 (33.8%) of patients had proctitis at diagnosis. Other symptoms were less frequent. The median severity score was 8 (IQR 5.8 to 10, range 1-17) out of 21. Fifteen (15/28, 53.6%) patients reported being vaccinated with MVA-BN before developing mpox.

All 130 MVA-BN-vaccinees born after 1976 as well as 66 (82.5%) of the 80 participants born before or in 1976 received two doses (**Table 1**). The remaining 14 vaccinees received one dose. Among those who received two doses, 59.7% (n=117/196) received both doses ID, 24.0% (ID-ID, n=47/196) received a mixed schedule (ID-SC, n=39/196, 19.9%; SC-ID, n=8/196, 4.1%), and 15.8% (n=32/196) received both doses SC (SC-SC).

### Long-term clinical outcomes of mpox

Almost all (186/196, 94.9%) mpox patients had skin or mucosal lesions at diagnosis (**Figure 2A-B-F and Supplement p5**). During evaluation at months 8, 16 and 24, 46.5% (33/71), 29.8% (17/57) and 31.7% (20/63) had visible scars (**Supplement p11**). While initial mpox skin lesions were painful in 38.8% (73/188), residual scars were associated with pain in only 2/71 participants at month 8, 1/57 at month 16, and 0/63 at month 24 (**Figure 2C**). The presence of scars 24 months after diagnosis was not significantly associated with mpox severity score (OR 1.06 [95% CI 0.86 to 1.32], *p*=.57), being born after 1976 (OR 0.71 [95% CI 0.18 to 2.86], *p*=.62), or HIV infection (OR 0.42 [95% CI 0.09 to 1.64], *p*=.23). Scars had little to no impact on daily life in most participants (DLQI score >5 in n=2 at month 16, and n=2 at month 24).

**Figure 2:**
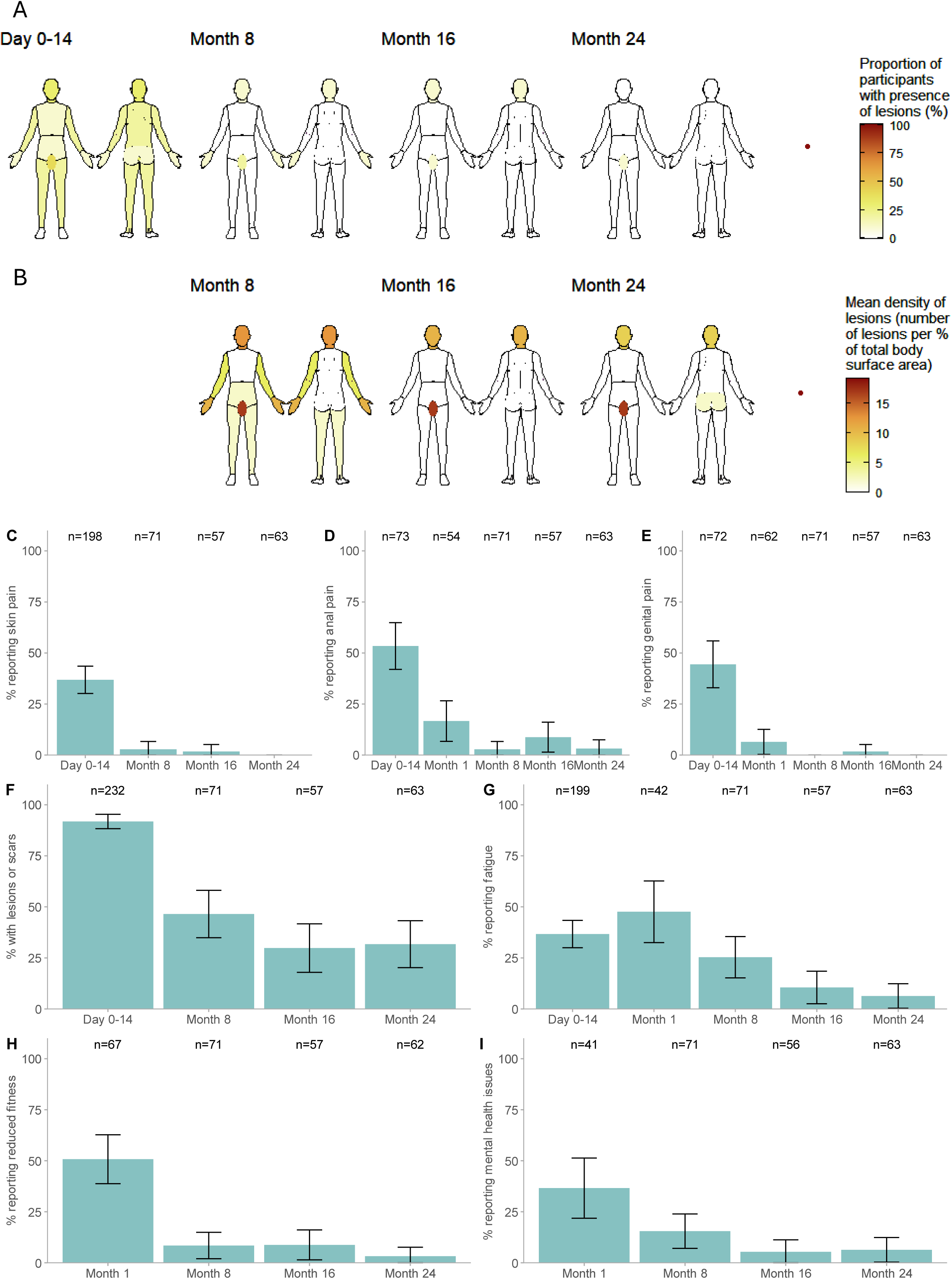
Long-term consequences of mpox. (A-B, F) Mpox-associated skin and mucosal lesions or scars at diagnosis, and 8 months, 16 months and 24 months after diagnosis; (A) proportion of participants with lesions or scars, per body area; (B) density of lesions or scars per body area, proportionate to body surface area (note: the self-reported presence or absence of anorectal scars was not verified by the investigator); (C-D-E) proportion of participants with pain at skin lesions or scars, in the anal area, or genital area (G) proportion of participants experiencing new-onset or worsened fatigue since mpox; (H) proportion of participants experiencing new-onset or worsened reduction in physical fitness since mpox; (I) proportion of participants experiencing new-onset or worsened mental health issues since mpox.

While half (39/73, 53.4%) of the patients had anal pain during mpox, this decreased to 16.7% (9/54) at month 1, 2.8% at month 8, 8.8% at month 16, and 3.2% at month 24 (**Figure 2D, Supplement p5**). During mpox, 44.4% (32/72) reported genital pain (**Figure 2E**). This decreased to 6.5% (4/62) at month 1, and later decreased further to <2%.

One third (73/189, 38.6%) of patients felt fatigued during mpox, 47.6% (20/42) at month 1, 25.4% (18/71) at month 8, 10.5% (6/57) at month 16, and 6.3% (4/63) at month 24 (**Figure 2G, Supplement p5**). Similarly, 50.7% (34/67) of patients expressed a reduction in physical fitness linked to mpox at month 1, and this proportion decreased to 3.2% (2/62) by month 24 (**Figure 2H**). The proportion of participants reporting new mental health problems linked to mpox followed similar trends (**Figure 2I**).

### Long-term virological outcomes after mpox

MPXV-PCR was negative on all samples from mpox patients 8-24 months post-infection (saliva and anorectal swabs: n=69, n=51, and n=63, each, at month 8, month 16, and month 24, respectively; semen: n=23 at month 8).

### Antibody dynamics

At baseline (pre-mpox and pre-MVA-BN), participants born before or in 1976 had 4.63-fold (95% CI 3.32-6.46, *p*<.001) higher VACV-lysate GMTs (**Figure 3A**), 2.93-fold (95% CI 2.15-4.00, *p*<.001) higher MPXV-E8 levels (**Figure 3B**), and a non-significant 3.90-fold (95% CI 0.97-15.71, *p*=.056) higher prevalence of MPXV-neutralising antibodies than participants born after 1976 (**Figure 3C**).

**Figure 3:**
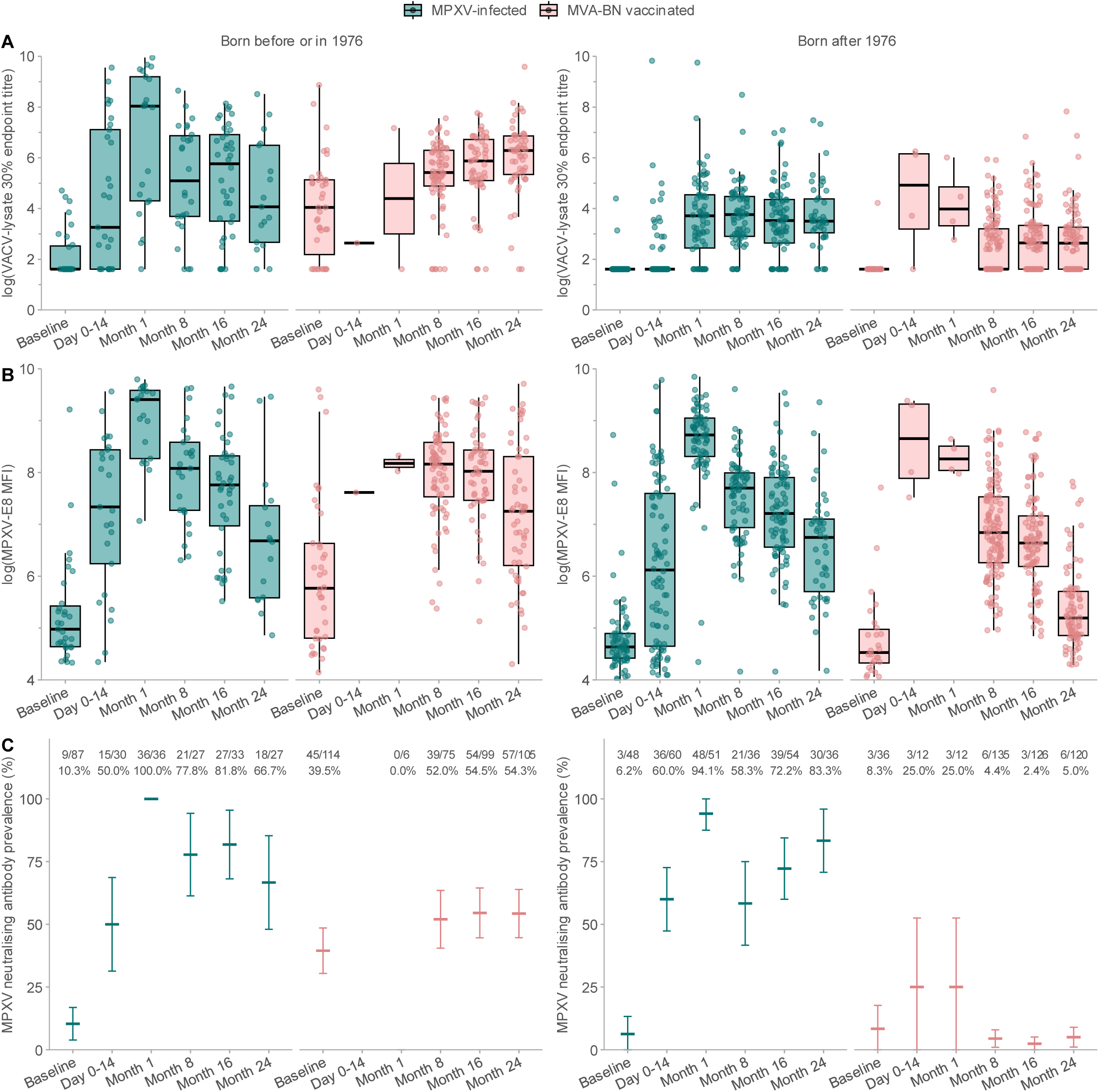
Antibody dynamics over time. (A) 30% endpoint titre of vaccinia virus (VACV)-lysate binding IgG measured by ELISA; (B) mpox virus (MPXV)-E8 binding IgG Mean Fluorescence Intensity (MFI) measured by bead-based Luminex assay; (C) MPXV-neutralising antibody prevalence with 95% confidence intervals. MVA-BN = modified vaccinia virus Ankara – Bavarian Nordic

**Figure 4:**
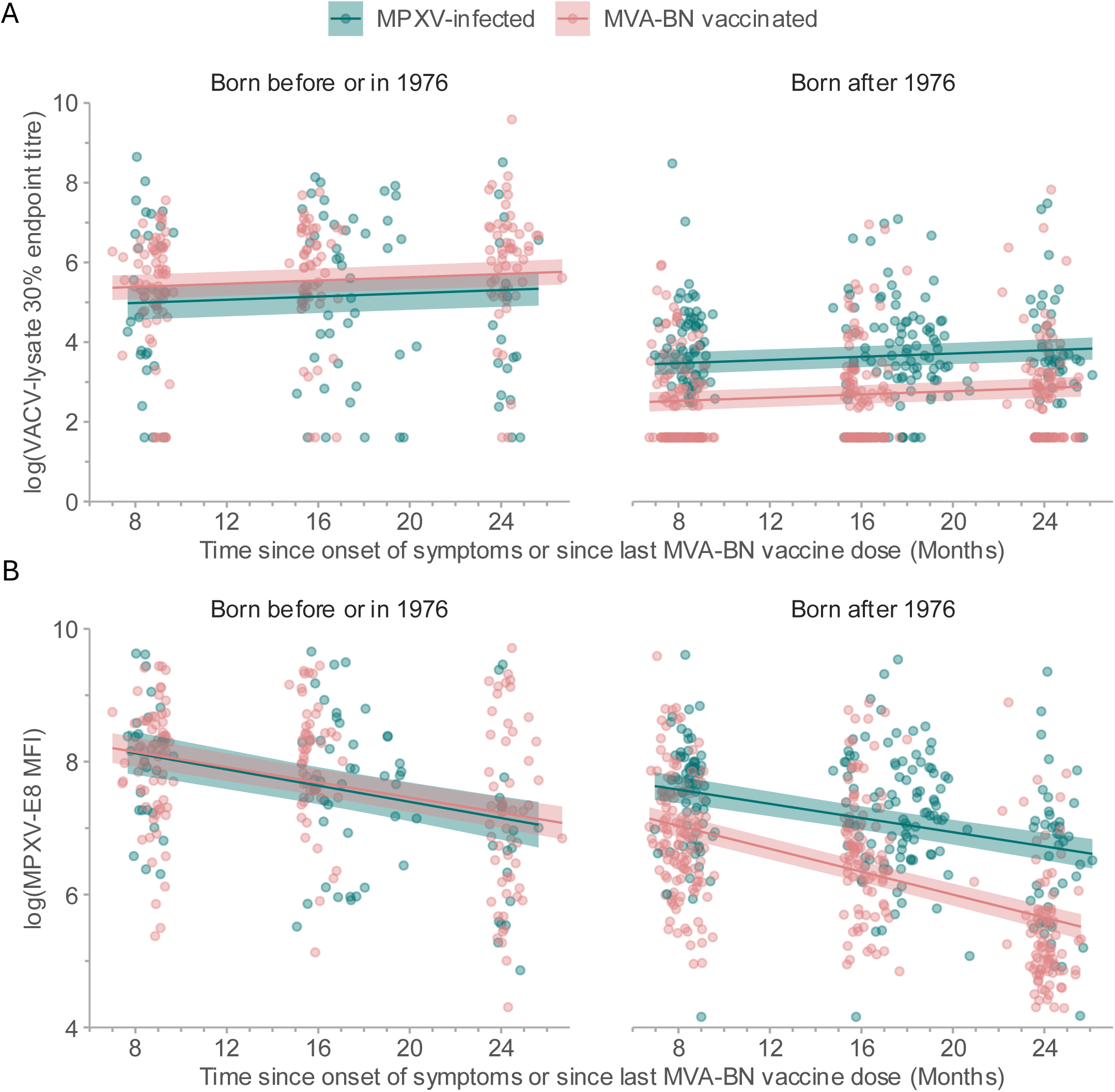
Association between binding antibody levels and exposure status (mpox virus [MPXV]-infection vs modified vaccinia virus Ankara – Bavarian Nordic [MVA-BN] vaccination) at months 8 to 24: observed values and values predicted by linear mixed effects regression, with 95% confidence interval (shaded area). (A) 30% endpoint titre of vaccinia virus (VACV)-lysate binding IgG measured by ELISA; (B) MPXV-E8 binding IgG Mean Fluorescence Intensity (MFI) measured by bead-based Luminex assay.

In a nested case control study comparing MPXV-infected individuals (cases) and MVA-BN vaccinees (controls) born before or in 1976, higher binding antibody titres were associated with protection against infection (OR 0.40 [95% CI 0.24 – 0.61], *p*<.001 for VACV-lysate antibodies; OR 0.47 [95% CI 0.25 – 0.77], *p* = .007 for MPXV-E8 antibodies). Likewise, the presence of MPXV-neutralising antibodies was associated with protection against infection (OR 0.18 [95% CI 0.04-0.62], *p*=.01)

Comparing VACV-lysate binding antibodies between sub-cohorts at month 8, we found the highest antibody titers in MPXV-infected individuals born before or in 1976 (GMT = 169.91, 95% CI 112.29-257.09), and in MVA-BN vaccinees born before or in 1976 (GMT = 254.10, 95% CI 187.96-343.52, **Table 2**, **Figures 3A & 4A**). Compared to MPXV-infected individuals born before or in 1976, antibody titres were 0.22-fold (95% CI 0.14-0.36, *p*<.001) lower in MPXV-infected individuals born after 1976 (GMT 37.51, 95% CI 28.83-48.81). The lowest antibody titers were found in MVA-BN vaccinees born after 1976 (GMT 14.57, 95% CI 11.54-18.40), which were 0.09-fold (95% CI 0.05-0.14, *p*<.001) lower compared to MPXV-infected individuals born before or in 1976 (**Supplement p6**). In all sub-cohorts, antibody titres increased slightly and at the same rate towards month 24 (2% increase in GMT per month [1.02-fold, 95% CI 1.01–1.03], *p* <.001).

**Table 2:** Associations between binding antibody levels or neutralising antibody prevalence and sub-cohort, after eight to twenty-four months of follow-up.

| Outcome | Model | Predictor | N participants | N observations | Estimated value* at month 8 | Fold-change | 95% CI | p-value |
| --- | --- | --- | --- | --- | --- | --- | --- | --- |
| **VACV-lysate binding antibody geometric mean titre** | LMER | MPXV-infected, born ≤ 1976 | 42 | 686 | 169.91 (112.29-257.09) | *Ref.* | *NA* | *NA* |
|  |  | MVA-BN vaccinated, born ≤ 1976 | 77 | 182 | 254.10 (187.96-343.52) | 1.50 | 0.90-2.49 | 0.12 |
|  |  | MPXV-infected, born > 1976 | 104 | 214 | 37.51 (28.83-48.81) | **0.22** | **0.14-0.36** | **<0.001** |
|  |  | MVA-BN vaccinated, born > 1976 | 128 | 315 | 14.57 (11.54-18.40) | **0.09** | **0.05-0.14** | **<0.001** |
|  |  | Time (months) | NA | NA | NA | **1.02** | **1.01-1.03** | **<0.001** |
| **MPXV-E8 binding antibody geometric mean MFI** | LMER | MPXV-infected, born ≤ 1976 | 42 | 84 | 3657.28 (2606.82-5131.06) | *Ref.* | *NA* | *NA* |
|  |  | MVA-BN vaccinated, born ≤ 1976 | 77 | 180 | 3714.46 (2962.28-4657.63) | 1.02 | 0.68-1.53 | 0.94 |
|  |  | MPXV-infected, born > 1976 | 104 | 213 | 2092.58 (1696.48-2581.15) | **0.57** | **0.38-0.85** | **0.006** |
|  |  | MVA-BN vaccinated, born > 1976 | 128 | 309 | 1265.43 (1066.04-1502.10) | **0.35** | **0.24-0.51** | **<0.001** |
|  |  | Time (months) | NA | NA | NA | **0.94** | **0.92-0.96** | **<0.001** |
|  |  | Time: MPXV-infected, born > 1976 | NA | NA | NA | 1.01 | 0.98-1.03 | 0.54 |
|  |  | Time: MVA-BN vaccinated, born > 1976 | NA | NA | NA | **0.98** | **0.95-1.00** | **0.029** |
|  |  | Time: MVA-BN vaccinated, born ≤ 1976 | NA | NA | NA | 1.00 | 0.98-1.03 | 0.76 |
| **MPXV-neutralising antibody prevalence** | GLM-lb | MPXV-infected, born ≤ 1976 | 21 | 21 | 0.81 (0.66-1.00) | *Ref.* | *NA* | *NA* |
|  |  | MVA-BN vaccinated, born ≤ 1976 | 50 | 50 | 0.56 (0.44-0.72) | **0.69** | **0.50-0.95** | **0.025** |
|  |  | MPXV-infected, born > 1976 | 36 | 36 | 0.69 (0.56-0.86) | 0.86 | 0.64-1.16 | 0.31 |
|  |  | MVA-BN vaccinated, born > 1976 | 73 | 73 | 0.03 (0.01-0.11) | **0.03** | **0.01-0.13** | **<0.001** |

CI = confidence interval; GLMER-lb = Generalised Linear Model with log binomial distribution; MFI = Mean Fluorescence Intensity; MPXV= Monkeypox virus; MVA-BN = Modified Vaccinia Virus Ankara - Bavarian-Nordic; VACV = Vaccinia Virus Significant associations ( $p < .05$ ) are indicated in bold \*geometric mean titre for VACV-lysate antibodies, geometric mean MFI for MPXV-E8 antibodies, prevalence for MPXV-neutralising antibodies

Findings were similar for MPXV-E8 antibody levels (**Figures 3B & 4B**), with the exception that MPXV-E8 antibody levels decreased over time, with the most rapid decay rate among MVA-BN vaccinees born after 1976 (8.26% decrease per month, 95% CI 9.09 to 7.42%, **Table 3**)

**Table 3:** Estimated decay rates and half-life of binding antibody levels, by sub-cohort.

| Antibody | Sub-cohort | Decay rate, % change per month (95% CI) |
| --- | --- | --- |
| VACV-lysate binding antibody geometric mean titre | MPXV-infected born > 1976 | NA |
|  | MVA-BN vaccinated born > 1976 | NA |
|  | MPXV-infected born ≤ 1976 | NA |
|  | MVA-BN vaccinated born ≤ 1976 | NA |
| MPXV-E8 binding antibody geometric mean MFI | MPXV-infected born > 1976 | -5.19 (-6.39 to -3.98) |
|  | MVA-BN vaccinated born > 1976 | -8.26 (-9.09 to -7.42) |
|  | MPXV-infected born ≤ 1976 | -5.92 (-7.85 to -3.94) |
|  | MVA-BN vaccinated born ≤ 1976 | -5.57 (-6.72 to -4.40) |

CI = confidence interval; MFI = Mean Fluorescence Intensity; MPXV= Monkeypox virus; MVA-BN = Modified Vaccinia Virus Ankara - Bavarian-Nordic; NA = Not Applicable (no significant decrease in titre over time); VACV = Vaccinia Virus

MPXV-infected individuals had estimated MPXV-neutralising antibody prevalences of 81% (95% CI 66-100%) in those born before or in 1976, and 69% (95% CI 56-86%) among those born after 1976 (comparison between groups: *p*=.32, **Figure 3C**). MVA-BN vaccinees born before or in 1976 had an estimated prevalence of 56% (95% CI 44-72%), which is 0.69-fold (95% CI 0.50–0.95, *p*=.025) lower compared to MPXV-infected individuals born before or in 1976. MVA-BN vaccinees born after 1976 had the lowest estimated prevalence (3%, 95% CI 1-11%), which is 0.03-fold (95% CI 0.01-0.13, *p*<.001) lower compared to MPXV-infected individuals born before or in 1976. Presence of MPXV-neutralising antibodies changed over time (from present to absent or vice-versa) in only n=11 individuals. Hence, these individuals were excluded from the model and time was not considered in the model.

HIV infection was associated with 2.00-fold (95% CI 1.04–3.85) higher VACV-lysate antibody levels among MVA-BN vaccinated individuals born before or in 1976, with 1.49-fold (95% CI 1.06–2.09) higher MPXV-E8 antibody levels among MPXV-infected individuals born after 1976, and with a 5% slower (1.05 [95% CI 1.02– 1.07]) decay of MPXV-E8 antibodies over time among MVA-BN vaccinated individuals born before or in 1976 (**Supplement p7, p12**). The remaining associations with binding antibody levels or neutralising antibody prevalence were not statistically significant.

Mpox severity score was not associated with binding antibody levels or neutralising antibodies in any birth cohort (**Supplement p8**).

Among individuals born after 1976, ID-ID vaccination was associated with 0.26-fold (95% CI 0.17–0.40) lower VACV-lysate GMTs, and with 0.51-fold (95% CI 0.36–0.74) lower MPXV-E8 antibody levels compared to standard SC-SC vaccination (**Supplement p9, Supplement p13**). In addition, mixed vaccination routes were associated with 0.54-fold (95% CI 0.33–0.87) lower VACV-lysate GMTs (but not significantly lower MPXV-E8 antibody levels), compared to SC-SC vaccination. Antibody levels did not differ significantly by vaccination route among individuals born before or in 1976, and no significant associations of vaccination routes with neutralising antibody prevalence were identified across birth cohorts.

## Discussion and conclusions

We described the long-term complications of MPXV infection, evaluated the possibility of MPXV persistence, and used a complementary set of assays to compare antibody dynamics up to two years after MPXV infection and MVA-BN vaccination.

Scarring, mainly on the hands, face, or genitals, is the predominant long-term sequela, affecting about one-third of patients up to two years post-infection. Similar smaller studies found scars in 84% of 43 patients 4-6 months post-infection,^22^ in 45% of 61 patients 12-15 months post-infection,^20,21^ and in 62.5% of 40 patients 20-23 months post-infection.^23^ Although scarring is frequent, most patients report a low impact of scars on their quality of life.^22^ Symptoms of anogenital pain, fatigue and mpox-related mental health issues, often experienced during mpox, do not persist beyond 6-12 months.

We did not find indications that MPXV persists in the long term. This finding is in line with other studies, which have reported clearance of MPXV-DNA after two to three weeks in various samples, including saliva, stool,^26^ and up to two months in rectal swabs and semen.^27^

Antibody dynamics following MPXV-infection and MVA-BN vaccination strongly differ by prior smallpox vaccination status. Smallpox-vaccinated individuals often have detectable orthopoxvirus-specific antibodies prior to MPXV-infection or MVA-BN vaccination, which are subsequently boosted by either MPXV-infection or MVA-BN vaccination, resulting in comparably high antibody levels 8 to 24 months later. These findings corroborate previous reports demonstrating durable immunological memory following smallpox vaccination,^12,25^ and extend existing evidence by showing that a boosting effect remains observable up to two years following MPXV-infection or MVA-BN vaccination. Conversely, smallpox-naïve individuals have much lower antibody levels 8 to 24 months following MPXV-infection or MVA-BN vaccination, and the immunological impact of MVA-BN vaccination appears to be markedly less pronounced than that of MPXV-infection. Notably, MVA-BN vaccine recipients rarely have detectable neutralising antibodies 8 to 24 months post-vaccination, in contrast to most MPXV-infected individuals. These results are consistent with other studies, which reported waning neutralising antibody levels within the first year post-MVA-BN vaccination, despite the presence of circulating MPXV-specific memory B-cells.^13^ It is likely that such memory B-cells remain functionally competent, and allow for successful recall of neutralising antibody responses several years after primary vaccination, as demonstrated in one study.^14^ Nevertheless, our findings suggest that, in smallpox-naïve individuals, MPXV-infection induces more efficient B-cell maturation leading to a more robust and durable humoral immune response than MVA-BN vaccination. While it remains to be determined whether this translates into superior protection against (re-)infection with orthopoxviruses, the data indicate that MVA-BN vaccination does not mimic–let alone outperform–the immunogenicity of natural infection. This underscores the need for more effective mpox-specific vaccines capable of inducing long-lasting protective immunity.

Owing to vaccine shortages in 2022, several countries–including Belgium–implemented ID administration of MVA-BN instead of the SC route. This strategy was supported by a randomised trial showing that two ID doses of 0.1 mL were non-inferior to two SC doses of 0.5 mL in terms of immunogenicity within 208 days post-vaccination.^28^ However, our real-world observations challenge this conclusion in the longer term in smallpox-unvaccinated individuals. Specifically, we observed that two ID doses of MVA-BN were associated with lower circulating binding antibody levels compared to two SC doses–a difference that, to our knowledge, has not been previously reported. Although differences in antibody levels between vaccination routes may not substantially affect short-term vaccine effectiveness, the role of cellular and tissue-resident immunity, especially given the likely mucocutaneous route of MPXV entry, warrants further investigation, as long-term protection data remain limited due to the decline of clade IIb MPXV circulation since 2023.

In low-incidence settings, a correlate of protection would offer a valuable alternative to assess vaccine-induced immunity. Our study uniquely included both pre-MVA-BN and pre-mpox samples, enabling assessment of baseline immunity and infection risk; we found a negative association between antibody levels and MPXV infection. This relationship may be biased by unmeasured confounders such as behavioural changes or unreported infections. Nonetheless, our findings support the potential role of antibodies as correlates of protection.^29^ Prospective studies with baseline samples and detailed immunological profiling of reinfections and breakthrough cases are needed.

This study offers unique insights into multiple facets of MPXV infection: long-term clinical sequelae, viral persistence, and humoral immunity, as well as immune responses to MVA-BN vaccination. Using longitudinal data from a large cohort of infected and vaccinated individuals followed for two years, we examined the durability and kinetics of humoral immunity in a real-world context. Methodological challenges included variable follow-up schedules, which we addressed using advanced statistical models. The lack of fully validated serological assays was mitigated through complementary testing approaches. Additionally, smallpox vaccination status could not be directly confirmed; instead, we used year of birth relative to the cessation of routine smallpox vaccination in Belgium as a widely accepted proxy.^8,10,11,13^

We observed a marked decline in antibody levels over time, particularly among smallpox-naïve MVA-BN vaccinees, while infection-induced responses (especially neutralising antibodies) were more sustained. As breakthrough infections continue to emerge, these findings raise concerns about waning population-level immunity. Although early estimates of MVA-BN effectiveness ranged between 66% and 90%,^30^ long-term protection remains unstudied. Future studies should clarify whether declining antibodies reflect reduced protection and whether booster doses can restore or prolong immunity

## Data Availability

De-identified participant data collected for the study will be made available from the corresponding author upon reasonable request (i.e., when ethically viable without violating the protection of participants or other valid ethical, privacy, or security concerns).

## Acknowledgments

During the preparation of this work, the authors used ChatGPT-4o as an editing tool (prompt: “improve grammar and spelling”). After using this tool, the authors reviewed and edited the content as needed and take full responsibility for the content of the publication.

## Funding

This work was funded by the Research Foundation–Flanders (FWO, grant number G096222N to EBo, LL, JM, G069725N to LL, WA, KV, NBR, AT, CVD, and 12B1M24N to CVD) and by the Netherlands Organization for Health Research and Development (ZonMw, grant agreement 10150022310035) to LMZ and RDdV. EBa, EDV, and IB are members of the Institute of Tropical Medicine’s Outbreak Research Team, financially supported by the Department of Economy, Science, and Innovation of the Flemish government (EWI).

## Conflict of interest statement

LL has received institutional consultancy fees from BioNTech and institutional research funding from Sanofi; both not relevant for this work. All other authors declare no conflict of interest.

## Author contributions

Conceptualized the study and wrote the protocol: NBR, CVD, LL. Participant inclusion, investigation and data collection: CVD, NBR, SB, CC, EB, NDC, BS, LV, IB, PS, EBo, CK, JVG, LL. Conducted sample processing and analysis: JV, JC, FV, EW, OG, AVH, JW, LMZ. Conducted data analysis: CVD, NBR, JM, CK, AT, LMZ. Data management: CVD, NBR, JM, LMZ. Secured funding: LL, JM, Ebo, WA, KV, CVD, LMZ, RDdV. Supervision study proceedings: LL, JM, RDdV. Supervised laboratory procedures: JM, KKA, WA, RDdV, MVE, KV. Writing of the initial manuscript: CVD, NBR, JM. Reviewed the manuscript: all authors. Had full access to the data: LL, CVD, NBR, JM, CK, AT, LMZ, RDdV. All authors contributed to review and approved the final version of the manuscript.

## Notes

### Author Declarations

The study was approved by the Institutional Review Board of the Institute of Tropical Medicine Antwerp (references 1628/22 and 1641/22) and the Ethics Committee of the University Hospital of Antwerp (references 3797 and 4981).

