## Supplement for "Long-term clinical, virological and immunological consequences of mpox virus infection or modified vaccinia virus Ankara vaccination: a 24-month prospective cohort study"

### Supplement to Manuscript "Scarring, viral persistence, and antibody dynamics until two years after monkeypox virus infection and modified vaccinia Ankara vaccination: a cohort study"

### 1. Supplementary Methods

#### 1.1. Supplementary Methods 1: MPXV-PCR targeting the MPXV-TNF receptor gene

DNA was automatically extracted (Maxwell®, Promega, Madison, WI, USA; using 300 µl sample input and 75 µl elution volume). PCR was performed using PerfeCTa FastMix II PCR Reagents (Quantabio, Beverly, MA, USA) and MPXV generic primers/probe as described previously.[1,2] Amplification was carried out on a QuantStudio 5 instrument (ThermoFisher Scientific, Waltham, MA, USA). To save time and material, a pooled analysis protocol was validated in which samples (300 µL in total) from five patients were pooled per specimen type (saliva or anal Eswab), and each pool was mixed with 300 µL lysis buffer and 30 µL proteinase K for further analysis. The mix was inactivated for 20 minutes at 56 °C and further processed according to the manufacturer's instructions. The DNA was eluted in 75 µL buffer. Samples were tested using a generic MPXV PCR and an internal control. Internal validation showed that Ct values from pooled samples exceeded individual sample Ct values by 1.27 ± 1.83 (unpublished data). Semen samples were analysed separately. If no signal was measurable, the sample was considered PCR-negative. All samples weak signals were reanalysed and considered PCR-positive if the signal was confirmed during repeat testing.

#### 1.2. Supplementary Methods 2: XMAP Luminex bead-based IgG immunoassay

We procured commercially available recombinant protein E8 derived from monkeypox virus (MPXV). The lyophilized protein was resuspended in distilled water according to the manufacturer's instructions and stored appropriately until use. The antigen was coupled to a maximum of 1.25 × 10^6 paramagnetic MAGPLEX COOH-microspheres (Luminex Corporation, Austin, TX), as previously described.[3,4] The coupling process involved a concentration of 4 µg of antigen per 1 × 10^6 beads. Additionally, bovine serum albumin (BSA, Sigma-Aldrich, St. Louis, USA; 3 µg) and tetanus toxoid were coupled to a separate set of beads to serve as background controls.

Next, we added beads and diluted sera in PBS (1%BSA, 0.05% azide) (1:100) to each well, bringing the final volume to 50 µl. The plates were incubated at 37°C for 30 minutes on a plate shaker, followed by three washes with 200 µl/well of 0.05% PBS-Tween 20. Next, 50 µl of a 1:500 dilution of secondary antibody (R-phycoerythrin-conjugated AffiniPure F(ab′)2 fragment of goat anti-human IgG, Jackson Immuno Research Laboratories) in PBS-BN was added to each well. The plates were incubated for 30 minutes at 37°C on a plate shaker set to 400 rpm, again in the dark. Following incubation, the plates were washed three times with PBS-BN-Tween, and the beads were resuspended in 125 µl of PBS-BN. The results were read using a Luminex® 100/200 analyzer, with an acquisition volume of 50 µl per well, DD gate settings of 5000-25000, and the high PMT option disabled. Results were reported as crude median fluorescent intensities (MFI). We repeated all samples that contained high levels of BSA-coupled bead signal (defined as > 4* SD) or contained low levels of Tetanus toxin-coated bead signal, defined as > or < 4* SD. If the repeat sample was still outside these cutoffs, we removed the sample from further analysis. For assay validation, we included a negative control panel consisting of 117 presumably smallpox-vaccinated individuals (born < 1976) and 196 presumably smallpox-vaccine naive individuals (born ≥ 1976).

##### 1.3. References to Supplementary Methods

1 Li Y, Zhao H, Wilkins K, Hughes C, Damon IK. Real-time PCR assays for the specific detection of monkeypox virus West African and Congo Basin strain DNA. *Journal of Virological Methods* 2010; **169**: 223–7.

2 De Baetselier I, Van Dijck C, Kenyon C, *et al.* Retrospective detection of asymptomatic monkeypox virus infections among male sexual health clinic attendees in Belgium. *Nature Medicine* 2022; **28**: 2288–92.

3 Yates JL, Hunt DT, Kulas KE, *et al.* Development of a novel serological assay for the detection of mpox infection in vaccinated populations. *Journal of Medical Virology* 2023; **95**: e29134.

4 Mariën J, Ceulemans A, Michiels J, *et al.* Evaluating SARS-CoV-2 spike and nucleocapsid proteins as targets for antibody detection in severe and mild COVID-19 cases using a Luminex bead-based assay. *Journal of Virological Methods* 2021; **288**: 114025.

#### 2. Supplementary Tables

##### 2.1. Supplementary Table 1: Mpox severity score

| Feature | Number of points |
|---|---|
| **Number of lesions** | |
| 0-4 | 1 |
| 5-24 | 2 |
| 25-100 | 3 |
| >100 | 4 |
| **Number of affected body areas** | |
| None | 0 |
| 1 | 1 |
| 2 | 2 |
| 3 | 3 |
| 4-5 | 4 |
| >5 | 5 |
| **Proctitis** | |
| Present | 3 |
| Absent | 0 |
| **Urethritis** | |
| Present | 2 |
| Absent | 0 |
| **Tonsillitis or throat ache** | |
| Present | 1 |
| Absent | 0 |
| **Fever or chills** | |
| Present | 2 |
| Absent | 0 |
| **Lymph nodes (regional or generalised)** | |
| Present | 1 |
| Absent | 0 |
| **Arthralgia or myalgia** | |
| Present | 1 |
| Absent | 0 |
| **Headache** | |
| Present | 1 |
| Absent | 0 |
| **Fatigue** | |
| Present | 1 |
| Absent | 0 |
| **Total** | Range 1 - 21 |

##### 2.2. Supplementary Table 2: Long-term sequelae

| | During mpox (n= 199) | | Month 1 (n= 74) | | Month 8 (n= 71) | | Month 16 (n=57 ) | | Month 24 (n= 63) | |
|---|---|---|---|---|---|---|---|---|---|---|
| | **n/N (%)** | **95% CI** | **n/N (%)** | **95% CI** | **n/N (%)** | **95% CI** | **n/N (%)** | **95% CI** | **n/N (%)** | **95% CI** |
| **Presence of skin lesions or scars** | 186/196 (94.9) | 91.8 - 98 | NA | - | 33/71 (46.5) | 34.9 - 58.1 | 17/57 (29.8) | 17.9 - 41.7 | 20/63 (31.7) | 20.3 - 43.2 |
| **Pain from skin lesions/scars** | 73/188 (38.8) | 31.9 - 45.8 | NA | - | 2/71 (2.8) | 0 - 6.7 | 1/57 (1.8) | 0 - 5.2 | 0/63 (0) | 0 - 0 |
| **DLQI score** | NA | - | NA | - | NA | - | | | | |
| 0-1 (no/minimal) | | | | | | | 52/56 (92.9) | 86.1 - 99.6 | 57/63 (90.5) | 83.2 - 97.7 |
| 2-5 (mild) | | | | | | | 2/56 (3.6) | 0 - 8.4 | 4/63 (6.3) | 0.3 - 12.4 |
| 6-10 (moderate) | | | | | | | 2/56 (3.6) | 0 - 8.4 | 2/63 (3.2) | 0 - 7.5 |
| 11-20 (severe) | | | | | | | 0/56 (0) | 0 - 0 | 0/63 (0) | 0 - 0 |
| >20 (very severe) | | | | | | | 0/56 (0) | 0 - 0 | 0/63 (0) | 0 - 0 |
| **Anal lesions** | 67/187 (35.8) | 29 - 42.7 | NA | - | 0/71 (0) | 0 - 0 | 1/57 (1.8) | 0 - 5.2 | 1/63 (1.6) | 0 - 4.7 |
| **Anal pain** | 39/73 (53.4)* | 42 - 64.9 | 9/54 (16.7) | 6.7 - 26.6 | 2/71 (2.8) | 0 - 6.7 | 5/57 (8.8) | 1.4 - 16.1 | 2/63 (3.2) | 0 - 7.5 |
| **Genital lesions** | 97/187 (51.9) | 44.7 - 59 | NA | - | 14/71 (19.7) | 10.5 - 29 | 9/57 (15.8) | 6.3 - 25.3 | 12/63 (19) | 9.4 - 28.7 |
| **Genital pain** | 32/72 (44.4)* | 33 - 55.9 | 4/62 (6.5) | 0.3 - 12.6 | 0/71 (0) | 0 - 0 | 1/57 (1.8) | 0 - 5.2 | 0/63 (0) | 0 - 0 |
| **Fatigue** | 73/189 (38.6) | 31.7 - 45.6 | 20/42 (47.6) | 32.5 - 62.7 | 18/71 (25.4) | 15.2 - 35.5 | 6/57 (10.5) | 2.6 - 18.5 | 4/63 (6.3) | 0.3 - 12.4 |
| **Reduction in physical fitness** | NA | - | 34/67 (50.7) | 38.8 - 62.7 | 6/71 (8.5) | 2 - 14.9 | 5/57 (8.8) | 1.4 - 16.1 | 2/62 (3.2) | 0 - 7.6 |
| **New or worsened mental health problems related to mpox** | NA | - | 15/41 (36.6) | 21.8 - 51.3 | 11/71 (15.5) | 7.1 - 23.9 | 3/56 (5.4) | 0 - 11.3 | 4/63 (6.3) | 0.3 - 12.4 |

DLQI = Dermatology Life Quality Index

**2.3. Supplementary Table 3: Estimated fold-change in binding antibody levels or neutralising antibody prevalences between sub-cohorts at month 8, calculated based on the regression models in Table 2**

| Outcome | Model | Reference sub-cohort | | Compared sub-cohort | | Fold-change (95% CI) | *p*-value |
|---|---|---|---|---|---|---|---|
| | | **Infection vs vaccination** | **Birth cohort** | **Infection vs vaccination** | **Birth cohort** | | |
| **VACV-lysate antibody geometric mean titre** | LMER | MPXV-infected | > 1976 | MVA-BN vaccinated | > 1976 | **2.57 (1.62 – 4.09)** | **<0.001** |
| | | MPXV-infected | > 1976 | MPXV-infected | ≤ 1976 | **0.22 (0.12 – 0.42)** | **<0.001** |
| | | MPXV-infected | > 1976 | MVA-BN vaccinated | ≤ 1976 | **0.15 (0.09 – 0.25)** | **<0.001** |
| | | MVA-BN vaccinated | > 1976 | MPXV-infected | ≤ 1976 | **0.09 (0.05 – 0.16)** | **<0.001** |
| | | MVA-BN vaccinated | > 1976 | MVA-BN vaccinated | ≤ 1976 | **0.06 (0.04 – 0.10)** | **<0.001** |
| | | MPXV-infected | ≤ 1976 | MVA-BN vaccinated | ≤ 1976 | 0.67 (0.34 – 1.31) | 0.41 |
| **MPXV-E8 antibody geometric mean MFI** | LMER | MPXV-infected | > 1976 | MVA-BN vaccinated | > 1976 | **1.65 (1.16 – 2.36)** | **0.002** |
| | | MPXV-infected | > 1976 | MPXV-infected | ≤ 1976 | **0.57 (0.34 – 0.97)** | **0.031** |
| | | MPXV-infected | > 1976 | MVA-BN vaccinated | ≤ 1976 | **0.56 (0.38 – 0.85)** | **0.002** |
| | | MVA-BN vaccinated | > 1976 | MPXV-infected | ≤ 1976 | **0.35 (0.21 – 0.57)** | **<0.001** |
| | | MVA-BN vaccinated | > 1976 | MVA-BN vaccinated | ≤ 1976 | **0.34 (0.24 – 0.49)** | **<0.001** |
| | | MPXV-infected | ≤ 1976 | MVA-BN vaccinated | ≤ 1976 | 0.99 (0.58 – 1.68) | 0.99 |
| **MPXV-neutralising prevalence** | GLM-lb | MPXV-infected | > 1976 | MVA-BN vaccinated | > 1976 | **25.35 (4.14 – 155.38)** | **<0.001** |
| | | MPXV-infected | > 1976 | MPXV-infected | ≤ 1976 | 0.86 (0.58 – 1.27) | 0.75 |
| | | MPXV-infected | > 1976 | MVA-BN vaccinated | ≤ 1976 | 1.24 (0.81 – 1.91) | 0.571 |
| | | MVA-BN vaccinated | > 1976 | MPXV-infected | ≤ 1976 | **0.03 (0.01 – 0.21)** | **<0.001** |
| | | MVA-BN vaccinated | > 1976 | MVA-BN vaccinated | ≤ 1976 | **0.05 (0.01 – 0.30)** | **<0.001** |
| | | MPXV-infected | ≤ 1976 | MVA-BN vaccinated | ≤ 1976 | 1.45 (0.95 – 2.20) | 0.111 |

CI = confidence interval; GLMER-lb = Generalised Linear Model with log binomial distribution; MFI = Mean Fluorescence Intensity; MPXV= Monkeypox virus; MVA-BN = Modified Vaccinia Virus Ankara - Bavarian-Nordic; VACV = Vaccinia Virus; Significant associations ($p$ <.05) are indicated in bold

#### 2.4. Supplementary Table 4: Associations between binding antibody levels or neutralising antibody prevalence and HIV status, by sub-cohort, after eight to twenty-four months of follow-up

| Outcome | Model | Population included in the model (sub-cohort) | | Predictor | N participants | N observations | Fold-change | 95% CI | *p*-value |
|---|---|---|---|---|---|---|---|---|---|
| | | Exposure status | Birth cohort | | | | | | |
| **VACV-lysate binding antibody geometric mean titre** | LMER | MPXV-infected | ≤ 1976 | No HIV infection | 22 | 44 | *Ref.* | *NA* | NA |
| | | | | HIV infection | 19 | 41 | 0.57 | 0.15 – 2.14 | 0.40 |
| | | | | Time (months) | NA | NA | 1.00 | 0.98 – 1.03 | 0.67 |
| | LMER | MPXV-infected | > 1976 | No HIV infection | 60 | 127 | *Ref.* | *NA* | |
| | | | | HIV infection | 41 | 82 | 1.01 | 0.62 – 1.66 | 0.95 |
| | | | | Time (months) | NA | NA | 1.01 | 0.99 – 1.03 | 0.34 |
| | LMER | MVA-BN vaccinated | ≤ 1976 | No HIV infection | 52 | 119 | *Ref.* | *NA* | NA |
| | | | | HIV infection | 25 | 63 | **2.00** | **1.04 – 3.85** | **0.038** |
| | | | | Time (months) | NA | NA | **1.03** | **1.02 – 1.05** | **<0.001** |
| | LMER | MVA-BN vaccinated | > 1976 | No HIV infection | 103 | 252 | *Ref.* | *NA* | |
| | | | | HIV infection | 25 | 63 | 1.10 | 0.69 – 1.77 | 0.69 |
| | | | | Time (months) | NA | NA | **1.02** | **1.01 – 1.04** | **0.003** |
| **MPXV-E8 binding antibody geometric mean MFI** | LMER | MPXV-infected | ≤ 1976 | No HIV infection | 22 | 44 | *Ref.* | *NA* | *NA* |
| | | | | HIV infection | 19 | 39 | 0.98 | 0.51 – 1.89 | 0.96 |
| | | | | Time (months) | NA | NA | **0.94** | **0.93 – 0.96** | **<0.001** |
| | LMER | MPXV-infected | > 1976 | No HIV infection | 60 | 127 | *Ref.* | *NA* | *NA* |
| | | | | HIV infection | 41 | 82 | **1.49** | **1.06 – 2.09** | **0.021** |
| | | | | Time (months) | NA | NA | **0.95** | **0.94 – 0.96** | **<0.001** |
| | LMER | MVA-BN vaccinated | ≤ 1976 | No HIV infection | 52 | 119 | *Ref.* | *NA* | *NA* |
| | | | | HIV infection | 25 | 61 | 1.00 | 0.60 – 1.69 | 0.99 |
| | | | | Time (months) | NA | NA | **0.93** | **0.92 – 0.94** | **<0.001** |
| | | | | Time: HIV infection | NA | NA | **1.05** | **1.02 – 1.07** | **0.001** |
| | LMER | MVA-BN vaccinated | > 1976 | No HIV infection | 103 | 248 | *Ref.* | *NA* | *NA* |
| | | | | HIV infection | 25 | 61 | 1.19 | 0.82 – 1.72 | 0.36 |
| | | | | Time (months) | NA | NA | **0.92** | **0.91 – 0.93** | **<0.001** |
| **MPXV neutralising antibody prevalence** | GLM-lb | MPXV-infected | ≤ 1976 | No HIV infection | 12 | 12 | *Ref.* | *NA* | *NA* |
| | | | | HIV infection | 8 | 8 | 0.90 | 0.56-1.44 | 0.663 |
| | GLM-lb | MPXV-infected | > 1976 | No HIV infection | 20 | 20 | *Ref.* | *NA* | *NA* |
| | | | | HIV infection | 15 | 15 | 1.13 | 0.69-1.81 | 0.594 |
| | GLM-lb | MVA-BN vaccinated | ≤ 1976 | No HIV infection | 32 | 32 | *Ref.* | *NA* | *NA* |
| | | | | HIV infection | 18 | 18 | 0.84 | 0.45-1.40 | 0.536 |
| | GLM-lb | MVA-BN vaccinated | > 1976 | No HIV infection | 57 | 57 | *Ref.* | *NA* | *NA* |
| | | | | HIV infection | 16 | 16 | NA$ | *NA* | *NA* |

CI = confidence interval; LMER = Linear Mixed Effects Regression (including participant identifier as random intercept); MPXV= Monkeypox virus; MVA-BN = Modified Vaccinia Virus- Bavarian-Nordic; VACV = Vaccinia Virus; Significant associations (*p*<.05 are indicated in bold); $ none of the MVA-BN vaccinees born >1976 with HIV infection had MPXV-neutralising antibodies above the cutoff. The model was discarded because of unrealistic model estimates.

#### 2.5. Supplementary Table 5: Associations between binding antibody levels or neutralising antibody prevalence and mpox severity, by sub-cohort, after eight to twenty-four months of follow-up

| Outcome | Model | Population included in the model (sub-cohort) | | Predictor | N participants | N observations | Fold-change | 95% CI | *p*-value |
|---|---|---|---|---|---|---|---|---|---|
| | | Exposure status | Birth cohort | | | | | | |
| **VACV-lysate binding antibody geometric mean titre** | LMER | MPXV-infected | ≤ 1976 | Severity score | 34 | 70 | 0.81 | 0.63 – 1.05 | 0.11 |
| | | | | Time (months) | NA | NA | 1.01 | 0.99 – 1.04 | 0.39 |
| | LMER | MPXV-infected | > 1976 | Severity score | 79 | 167 | 1.02 | 0.94 – 1.11 | 0.58 |
| | | | | Time (months) | NA | NA | 1.00 | 0.98 – 1.02 | 0.95 |
| **MPXV-E8 binding antibody geometric mean MFI** | LMER | MPXV-infected | ≤ 1976 | Severity score | 34 | 68 | 0.97 | 0.86 – 1.11 | 0.68 |
| | | | | Time (months) | NA | NA | **0.94** | **0.92 – 0.96** | **<0.001** |
| | LMER | MPXV-infected | > 1976 | Severity score | 79 | 167 | 1.02 | 0.96 – 1.08 | 0.46 |
| | | | | Time (months) | NA | NA | **0.94** | **0.93 – 0.95** | **<0.001** |
| **MPXV neutralising antibody prevalence** | GLM-logistic | MPXV-infected | ≤ 1976 | Severity score | 18 | 18 | 0.50* | 0.13-1.02 | 0.169 |
| | GLM-logistic | MPXV-infected | > 1976 | Severity score | 26 | 26 | 1.24* | 0.89-1.84 | 0.234 |

CI = confidence interval; GLM-logistic = Generalised linear model (logistic regression); LMER = Linear Mixed Effects Regression (including participant identifier as random intercept); MPXV= Monkeypox virus; MVA-BN = Modified Vaccinia Virus- Bavarian-Nordic; VACV = Vaccinia Virus

Significant associations (*p*<.05 are indicated in bold)

*Odds Ratio

**2.6. Supplementary Table 6: Associations between binding antibody levels or neutralising antibody prevalence and MVA-BN vaccination route, by sub-cohort, after eight to twenty-four months of follow-up**

| Outcome | Model | Population included in the model (sub-cohort) | | Predictor | N participants | N observations | Fold-change | 95% CI | *p*-value |
|---|---|---|---|---|---|---|---|---|---|
| | | Exposure status | Birth cohort | | | | | | |
| **VACV-lysate binding antibody geometric mean titre** | LMER | MVA-BN vaccinated | ≤ 1976 | SC | 10 | 22 | *Ref.* | *NA* | *NA* |
| | | | | SC-SC | 5 | 10 | 0.93 | 0.19 – 4.41 | 0.92 |
| | | | | ID | 3 | 6 | 2.14 | 0.33 – 13.77 | 0.42 |
| | | | | ID-ID | 45 | 113 | 1.04 | 0.39 – 2.80 | 0.93 |
| | | | | Mixed | 14 | 31 | 0.95 | 0.30 – 3.08 | 0.94 |
| | | | | Time (months) | NA | NA | **1.03** | **1.02 – 1.05** | **<0.001** |
| | LMER | MVA-BN vaccinated | > 1976 | SC-SC | 27 | 66 | *Ref.* | *NA* | *NA* |
| | | | | ID-ID | 68 | 167 | **0.26** | **0.17 – 0.40** | **<0.001** |
| | | | | Mixed | 33 | 82 | **0.54** | **0.33 – 0.87** | **0.012** |
| | | | | Time (months) | NA | NA | **1.02** | **1.01 – 1.04** | **0.002** |
| **MPXV-E8 binding antibody geometric mean MFI** | LMER | MVA-BN vaccinated | ≤ 1976 | SC | 10 | 22 | *Ref.* | *NA* | *NA* |
| | | | | SC-SC | 5 | 10 | 0.68 | 0.22 – 2.05 | 0.49 |
| | | | | ID | 3 | 5 | 1.20 | 0.31-4.62 | 0.79 |
| | | | | ID-ID | 45 | 112 | 1.01 | 0.50 – 2.04 | 0.98 |
| | | | | Mixed | 14 | 31 | 0.86 | 0.37 – 1.97 | 0.71 |
| | | | | Time (months) | NA | NA | **0.94** | **0.93-0.96** | **<0.001** |
| | LMER | MVA-BN vaccinated | > 1976 | SC-SC | 27 | 66 | *Ref.* | *NA* | *NA* |
| | | | | ID-ID | 68 | 163 | **0.51** | **0.36 – 0.74** | **<0.001** |
| | | | | Mixed | 33 | 80 | 0.69 | 0.46 – 1.05 | 0.081 |
| | | | | Time (months) | NA | NA | **0.92** | **0.91 – 0.93** | **<0.001** |
| **MPXV neutralising antibody prevalence** | GLM-logistic | MVA-BN vaccinated | ≤ 1976 | SC | 8 | 8 | *Ref.* | *NA* | *NA* |
| | | | | SC SC | 4 | 4 | 0.80* | 0.16-2.21 | 0.70 |
| | | | | ID ID | 29 | 29 | 0.77* | 0.42-1.77 | 0.44 |
| | | | | ID | 1** | 1** | NA | *NA* | *NA* |
| | | | | Mixed | 8 | 8 | 1.20* | 0.58-2.74 | 0.59 |
| | GLM- logistic | MVA-BN vaccinated | > 1976 | SC SC | 16 | 16 | NA$ | *NA* | *NA* |
| | | | | ID ID | 32 | 32 | NA$ | *NA* | *NA* |
| | | | | Mixed | 25 | 25 | NA$ | *NA* | *NA* |

CI = confidence interval; GLM-lb = Generalised Linear Model with log binomial distribution; GLM-logistic = Generalised linear model (logistic regression); ID = one intradermal vaccine dose; ID-ID = two intradermal vaccine doses; LMER = Linear Mixed Effects Regression (including participant identifier as random intercept); Mixed = one intradermal and one subcutaneous dose, or vice-versa; MPXV= Monkeypox virus; MVA-BN = Modified Vaccinia Virus- Bavarian-Nordic; NA = Not Applicable; SC = one subcutaneous vaccine dose; SC-SC = two subcutaneous vaccine doses; VACV = Vaccinia Virus

Significant associations ($p$<.05 are indicated in bold); *Odds Ratio; **only one observation, which was therefore omitted from the model; $ only two MVA-BN vaccinated individuals born >1976 had MPXV-neutralising antibodies above the cutoff. Both of them received their vaccines twice intradermally. The model was discarded because of unrealistic model estimates.

#### 3. Supplementary Figures

##### 3.1. Supplementary Figure 1: Association between binding antibody levels at months 8 to 24 and HIV infection

(A-H) Observed values and values predicted by linear mixed effects regression, with 95% confidence interval (shaded area). (A-D) 30% endpoint titre of VACV-lysate binding IgG measured by ELISA; (E-H) MPXV-E8 binding IgG Mean Fluorescence Intensity measured by bead-based Luminex assay; Data were modeled separately by birth cohort (born before or in 1976 vs after 1976) and exposure status (MPXV-infection vs MVA-BN vaccination).

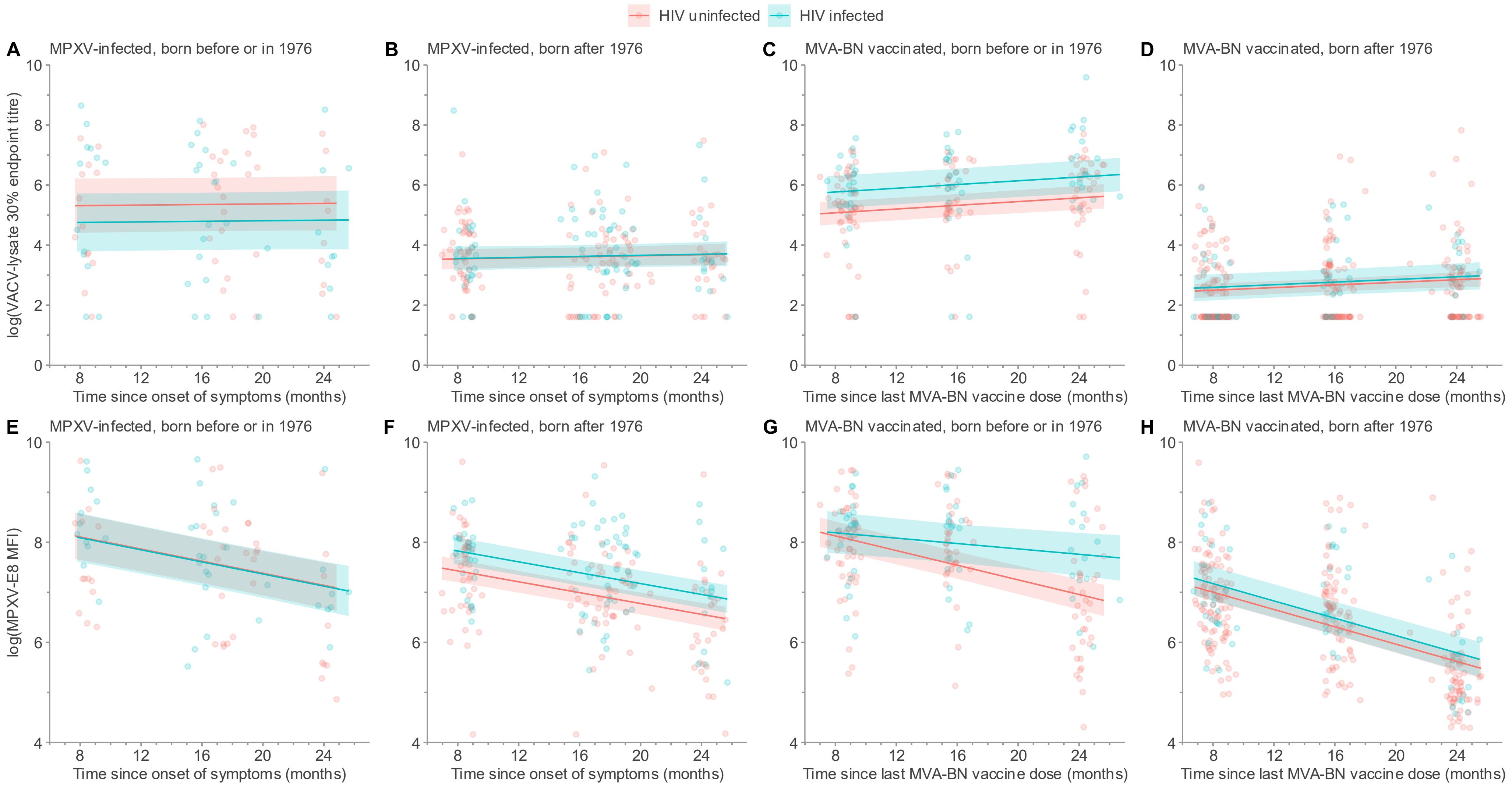

##### 3.2. Supplementary Figure 2: Association between binding antibody levels at months 8 to 24 and MVA-BN vaccination route

(A-D) Observed values and values predicted by linear mixed effects regression, with 95% confidence interval (shaded area). (A-B) 30% endpoint titre of VACV-lysate binding IgG measured by ELISA; (C-D) MPXV-E8 binding IgG Mean Fluorescence Intensity measured by bead-based Luminex assay; Data were modeled separately by birth cohort (born before or in 1976 vs after 1976). ID: intradermal; SC: subcutaneous

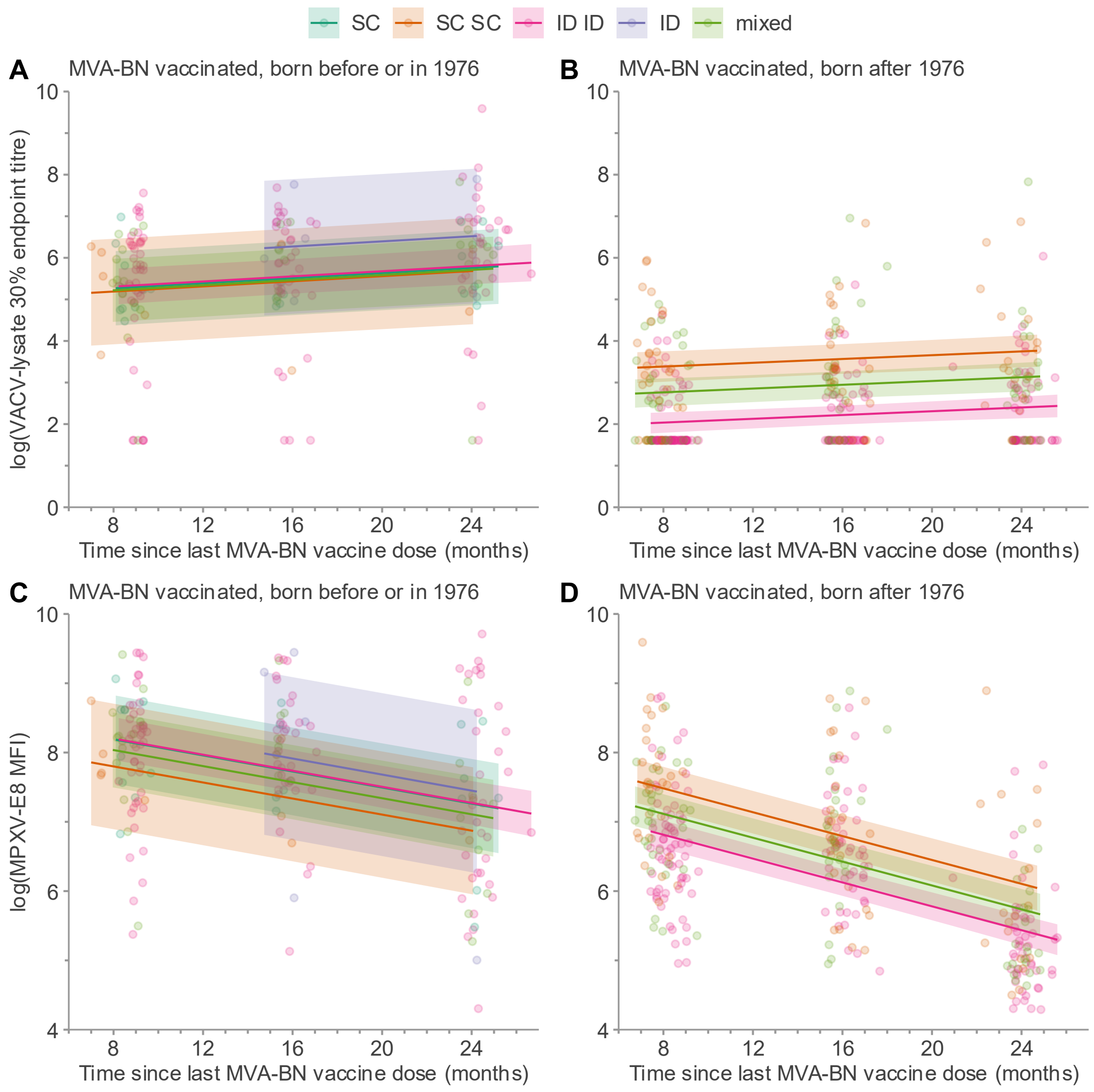
